# International healthcare workers’ experiences and perceptions of the 2022 multi-country mpox outbreak

**DOI:** 10.1101/2024.08.23.24312504

**Authors:** Vanessa Apea, Boghuma K. Titanji, Francesca H. Dakin, Rosalie Hayes, Melanie Smuk, Habiba Kawu, Laura Waters, Itsik Levy, Daniel R. Kuritzkes, Monica Gandhi, Jürgen Rockstroh, Mauro Schechter, Martin Holt, Romain Palich, Claudia P. Cortes, Silvia Nozza, Cristina Mussini, Aleaxandra Calmy, Brenda E. Crabtree Ramirez, José L. Blanco, Sanjay Bhagani, Claire Dewsnap, Chloe Orkin, the Mpox SHARE-NET writing group

**Affiliations:** SHARE Collaborative, Queen Mary University of London, London, United Kingdom; Blizard Institute, Queen Mary University of London, London, United Kingdom; Barts Health NHS Trust, Department of Infection and Immunity, London, United Kingdom; Emory University School of Medicine, Division of Infectious Diseases, Atlanta, United States; Atlanta Veterans Affairs Medical Center, Decatur, United States; Nuffield Department of Primary Care Health Sciences, University of Oxford, Oxford, United Kingdom; Wolfson Institute of Population Health, Queen Mary University of London, London, United Kingdom; University College London Hospitals NHS Foundation Trust, London, United Kingdom; Sheba Medical Center, Infectious Disease Unit, Ramat Gan, Israel; Brigham and Women’s Hospital, Harvard Medical School, Boston, United States; University of California, Department of Medicine, San Francisco, United States; University of Bonn, Bonn, Germany; Universidade Federal do Rio de Janeiro, Departamento de Doenças Infecciosas e Parasitárias, Rio de Janeiro, Brazil; Centre for Social Research in Health, University of New South Wales, Sydney, Australia; Pitié-Salpêtrière Hospital, Tropical Medicine and Infectious Disease Department, Paris, France; Universidad de Chile, Faculty of Medicine, Santiago, Chile; S. Raffaele University Hospital IRCCS, Division of Infectious Diseases, Milan, Italy; University of Modena and Reggio Emilia, Modena, Italy; Geneva University Hospitals, Division of Infectious Diseases, HIV/AIDS Unit, Geneva, Switzerland; Departamento de Infectología. Instituto Nacional de Ciencias Médicas y Nutrición, Mexico City, Mexico; University of Barcelona, Infectious Diseases Service, Hospital Clinic-IDIBAPS, Barcelona, Spain; Royal Free London NHS Foundation Trust, HIV services, Royal Free Hospital, London, United Kingdom; Sheffield Teaching Hospital NHS Foundation Trust, Sexual Health, Sheffield, United Kingdom

## Abstract

In May 2022, the most widespread outbreak of sustained transmission of mpox outside of countries historically affected countries in Western and Central Africa occurred. We aimed to examine the personal and clinical experiences of international healthcare workers (HCWs) during this public health emergency. We conducted an international cross-sectional survey study between August and October 2022, examining the experiences and perceptions of HCWs clinically involved with the 2022 mpox response. Respondents were recruited via an international network of sexual health and HIV clinicians responding to mpox and promoted through clinical associations and social media. Survey domains included: clinical workload; preparedness; training and support at work; psychological well-being and vaccination. 725 multi-national healthcare workers across 41 countries were included in the analysis. 91% were physicians specialised in Sexual Health or Infectious Diseases; with 34% (n=247) of all respondents involved in mpox policy. A substantial proportion of respondents (n=296, 41%) reported working longer hours during the mpox outbreak, with no concomitant removal of other clinical responsibilities. 30% (n=218) of respondents reported that they had never heard of mpox before the outbreak and over 25% of the respondents reported that they had misdiagnosed someone initially. This culminated in a high prevalence of moral distress at 30%. Less than 9% of HCWs in the region of the Caribbean, Central America and South America had been offered a vaccine as compared to almost 1/3 in the other regions. Where offered, there were high levels of uptake across all regions. Our findings highlight a critical need for addressing the profound gaps in HCW knowledge about re-emerging diseases with pandemic potential. Strengthening the resilience of global health systems and prioritising internationally coordinated approaches to global vaccine deployment is imperative.

## Introduction

Healthcare workers (HCWs) are essential pillars of health system preparedness and resilience in public health emergencies [1, 2]. The COVID-19 pandemic was a stark warning of the significant detriment that pandemics can inflict on the health and wellbeing of HCWs [3–5], and the response called for rapid and extensive transformation of already strained global health systems. At the population health level, entrenched structural health inequalities converged with transmission dynamics of SARS CoV-2. The confluence of these factors exposed system vulnerabilities and severely depleted staff capacity and infrastructure [4, 6]. Frontline HCWs were challenged with providing care and comfort to very sick people with a disease with a hitherto unknown clinical course and prognostic factors, alongside coping with uncertainty, contagion anxiety, fear of transmission to family networks, scarcity of personal protective equipment and extended working hours. This exerted a significant and pervasive toll on physical and psychological well-being of HCWs [4, 7, 8].

Reports on HCW experiences describe wide-ranging experiences of adversity, moral distress (the inability to provide appropriate care and act in line with personal and professional values) and resilience. In a British Medical Association survey of UK doctors after the second wave of COVID-19, 78% of respondents stated that ‘moral distress’ resonated with their experiences at work [4]. International knowledge exchange and transfer were identified as key to partially combat the difficulties experienced by HCWs during the COVID-19 pandemic.

It was within this context, that the 2022-23 multi-country outbreak of mpox occurred. Mpox (formerly known as monkeypox) is a disease caused by the zoonotic orthopoxvirus, human monkeypox virus (MPXV), which is endemic in the rainforest regions of several Central and West African countries [9–11]. In May 2022, simultaneous human mpox outbreaks began in Europe and extended across various countries that were not typically affected; occuring in the absence of epidemiological links to historically affected countries or infected animals [12]. Mpox was declared a public health emergency of international concern (PHEIC) by the World Health Organisation (WHO) two months later, reflecting the pace of global proliferation and the burden of disease attributable to the virus [2].

Mpox has been endemic in Central and Western Africa for over 50 years, but received scant global attention prior to the 2022 outbreak. This is despite repeated calls by African physicians and scientists for heightened prioritisation and investment into surveillance programmes, research and development to understand the evolution and epidemic potential of mpox in Africa [9, 13]. The 2022 global outbreak thus took many countries, and their health systems, by surprise. Most HCWs outside of historically affected countries had limited or no experience or knowledge of mpox [14]. The 2022 outbreak began with cases being predominantly described among sexual networks of men who have sex with men through close sexual contact with lesions and body fluids [15–17]. Clinical manifestations included prominent genital symptoms, including anogenital and mucosal lesions [15–17]. This differs markedly from traditional descriptions of zoonotically acquired cases. Due to this specific pattern of clinical symptoms, many patients were referred to Sexual Health Services where HCWs may have less experience of managing tropical disease. However, this sector does have extensive experience in epidemic management including the HIV epidemic.

Control measures involve isolating affected individuals, providing supportive care, and implementing public health measures such as vaccination campaigns and contact tracing [18, 19]. Both therapeutics for severe cases (e.g. tecovirimat) and vaccines used as mpox post-exposure prophylaxis or pre-exposure prophylaxis had originally been developed to treat the closely related smallpox (variola) virus [20, 21]. While these vaccinations and treatments did exist there were significant shortages even in high-income countries and as with COVID-19 an unacceptable and ongoing lack of access in resource-limited regions [22].

Mpox serves as a glaring reminder of the increasing risk posed by emerging pathogens and emergence of known pathogens in new contexts, and further underscores the importance of robust preparedness of infrastructure at global, regional and service levels. Mpox has placed a significant burden on healthcare systems and on HCWs still reeling from the impact of COVID-19 and pandemic fatigue. Understanding HCWs’ experience and needs during outbreaks is critical to strengthening health system resilience and developing effective epidemic/pandemic preparedness and response strategies [1]. Yet, literature describing HCWs’ experiences of the 2022 mpox outbreak is limited. The experiences of HCWs are often inferred indirectly from descriptions of the challenging environments in which they work. These environments are characterised by a complex mix of evolving and, sometimes conflicting policies and guidelines within healthcare systems already strained by the COVID-19 pandemic [23, 24].

Our study aimed to directly examine the personal and clinical experiences of international HCWs during the mpox outbreak, including their clinical management of mpox and their perceived sense of preparedness.

## Methods

We conducted an international cross-sectional survey study between 10^th^ August and 31^st^ October 2022, examining the experiences and perceptions of HCWs clinically involved with the 2022 mpox response. The survey was disseminated through clinical networks outside of previously affected regions. This analysis was restricted to individuals residing in the United Kingdom (UK), the European Union (EU), the Caribbean, Central America, South America, the United States (US) and Canada because the purpose of the survey was to evaluate pandemic preparedness and the clinical confidence of clinicians dealing with an existing pathogen emerging in a new context.

### Data collection

Anonymised, self-reported data was collected via an online survey containing 87 closed and open-text questions exploring: demographic characteristics; involvement in mpox clinical, research, and policy-related work; self-assessment of knowledge and confidence around mpox diagnosis and management and views on outbreak preparedness; educational resources; assessment of risk; workload; safety at work; vaccination; and perceptions and experiences of moral distress and injury. Moral distress was defined as the psychological unease generated where professionals identify an ethically correct action to take but are constrained in their ability to take that action. Moral injury was defined as arising where sustained moral distress leads to impaired function or longer-term psychological harm.

The survey was carried out in English, Spanish, French and Portuguese via the international collaboration SHARE-Net, an informal network established and led by academic researchers within the London-based SHARE Research Collaborative [26]. The survey was disseminated through newsletters and social media feeds of the British Association for Sexual Health and HIV, the British HIV Association, the European AIDS Clinical Society, the International AIDS Society, the Infectious Diseases Society of America and the research networks of SHARE-net collaborators from 16 countries. All survey questions examined in this analysis are availalable online at https://osf.io/jf6kw.

### Ethical approval and regulations

The survey was administered via a survey platform compliant with general data protection regulations (SMART Survey LTD, Tewkesbury, UK) and received ethical approval from the Queen Mary University of London Ethics of Research Committee (QMERC22.297). The opening page contained information about the aims of the study and custodianship and use of study data. The survey was piloted with ten sexual health clinicians in the UK. By clicking ‘continue’ and commencing the survey, individuals were considered to have given consent.

### Statistical analysis

We present descriptive statistics of frequency and percentages created for relevant questions. The response rate (denominator) is clearly noted. Partial responses were excluded from the analysis. Data remains in the original categories. Ethnicity was defined using nine categories, including a free text category. Three regional subgroups were created for our analyses; the first grouped European countries (including the EU and the UK), the second grouped Caribbean, Central America and South America countries and the third grouped the United States of America (USA) and Canada together. We examined demographic characteristics, impact on workload, prior knowledge, confidence and misdiagnosis, training and support received, vaccine uptake, moral distress and moral injury. We assessed differences by geographical location, drawing comparisons across the three regions. All analyses were performed using Stata Statistical Software: Release 17 (College Station, TX: StataCorp LLC). Results are presented as frequency (percentage): n (%).

## Results

### Respondent characteristics

A total of 725 respondents completed the survey across the regions of Europe (n=529), the Caribbean, Central America and South America (CCS) (n=114) and the United States of America (USA) and Canada (n=82). The demographic of respondents are summarised in Table 1. The analysis excluded an additional 48 respondents Of these, 36 were located outside of these regions across the WHO African region (n=18), South East Asian (n=2), Western Pacific (n=16) regions. 12 of those excluded did not provide their location.

**Table 1.** Summary of Characteristics.

|  | Europe N=529 | Caribbean, Central<br>America and<br>South America<br>(CCS) N=114 | United States<br>of America<br>(USA) and<br>Canada N=82 | Overall Total<br>N=725 |
| --- | --- | --- | --- | --- |
| **Age** | n=529 | n=114 | n=82 | n=725(100.00%) |
| 18-25 | 4(0.76%) | 0(0.00%) | 1(1.22%) | 5(0.69%) |
| 26-30 | 42(7.94%) | 15(13.16%) | 1(1.22%) | 58(8.00%) |
| 31-34 | 56(10.59%) | 12(10.53%) | 9(10.98%) | 77(10.62%) |
| 35-40 | 84(15.88%) | 17(14.91%) | 6(7.32%) | 107(14.76%) |
| 41-50 | 168(31.76%) | 29(25.44%) | 20(24.39%) | 217(29.93%) |
| 51-60 | 119(22.50%) | 13(11.40%) | 18(21.95%) | 150(20.69%) |
| 60+ | 56(10.59%) | 28(24.56%) | 27(32.93%) | 111(15.31%) |
| **Gender** | n=529 | n=114 | n=81 | n=724(99.86%) |
| Cis-Female | 302(57.09%) | 52(45.51%) | 42(51.85%) | 396(54.70%) |
| Cis-Male | 209(39.51%) | 59(51.75%) | 38(46.91%) | 306(42.27%) |
| Non-binary/non-conforming | 3(0.57%) | 1(0.88%) | 0(0.00%) | 4(0.55%) |
| Prefer not to say | 13(2.46%) | 1(0.88%) | 1(1.23%) | 15(2.07%) |
| Transfemale | 0(0.00%) | 1(0.88%) | 0(0.00%) | 1(0.14%) |
| Transmale | 2(0.38%) | 0(0.00%) | 0(0.00%) | 2(0.28%) |
| **Ethnicity** | n=514 | n=114 | n=82 | n=710(97.93%) |
| White | 467(90.86%) | 33(28.95%) | 56(68.29%) | 556(78.31%) |
| Black/African American | 8(1.56%) | 2(1.75%) | 6(7.32%) | 16(2.25%) |
| Asian/Asian American | 11(2.14%) | 3(2.63%) | 10(12.20%) | 24(3.38%) |
| Latinx or Hispanic | 6(1.17%) | 75(65.79%) | 5(6.10%) | 86(12.11%) |
| American Indian<br>or Alaska Native,<br>Middle Eastern or<br>North African,<br>Native Hawaiian,<br>Other Pacific<br>Islander, Mixed or<br>Multiple Ethnic<br>Group and Other<br>Ethnic Group. | 22(4.28%) | 1(0.88%) | 5(6.10%) | 28(3.94%) |

**Table 2.** Proportions of moral distress.

|  |  |  |  |
| --- | --- | --- | --- |
|  | Europe N=529 | Caribbean,<br>Central<br>America and<br>South America<br>N=114 | Canada and<br>USA N=82 |
| **Does the term “moral distress” resonate with your experiences at work managing suspected or confirmed clinical cases of mpox?**<br>**Definition of “moral distress”** - the psychological unease generated where professionals identify an ethically correct action to take but are constrained in their ability to take that action | n=523 | n=114 | n=81 |
| Does not resonate at all | 252 (48.18%) | 46 (40.35%) | 26 (32.10%) |
| Somewhat resonates | 80 (15.30%) | 23 (20.18%) | 13 (16.05%) |
| Slightly resonates | 112 (21.41%) | 22 (19.30%) | 26 (32.10%) |
| Moderately resonates | 43 (8.22%) | 13 (11.40%) | 9 (11.11%) |
| Extremely resonates | 36 (6.88%) | 10 (8.77%) | 7 (8.64%) |

**Table 3.** Proportions of moral injury.

|  |  |  |  |
| --- | --- | --- | --- |
| **Does the term “moral injury” resonate with your experiences at work managing suspected or confirmed clinical cases of mpox?** | n=520 | n=114 | n=81 |
| **Definition of “moral injury”** - sustained moral distress leading to impaired function or longer-term psychological harm |  |  |  |
| Does not resonate at all | 348 (66.92%) | 57 (50.00%) | 45 (55.56%) |
| Somewhat resonates | 47 (9.04%) | 20 (17.54%) | 10 (12.35%) |
| Slightly resonates | 90 (17.31%) | 21 (18.42%) | 17 (20.99%) |
| Moderately resonates | 19 (3.65%) | 9 (7.89%) | 4 (4.94%) |
| Extremely resonates | 16 (3.08%) | 7 (6.14%) | 5 (6.17%) |

Most were aged 41-50 years (n=217; 30%). Within the USA and Canada and Europe, 52% and 57% identified themselves as cis-female, respectively. Within the CCS region, the majority of respondents were cis male (n=59; 52%).

Those of white background formed most of the respondents in Europe (n=467; 91%) and the USA and Canada (n=56; 68%). Those of Latinx /Hispanic background formed the majority of those responding within the CCS region (n=75; 66%).

Overall the majority of respondents across all regions were physicians specialised in Sexual Health or Infectious Diseases (n=663; 91%); with 34% (n=247) of all respondents involved in mpox policy.

### Mpox related workload

A substantial proportion of respondents (n=296, 41%) reported working longer hours during the mpox outbreak. Within the USA and Canada, 60% (n=49) of respondents reported longer hours, as did 43% (n=225) of those based in Europe. This proportion was comparatively lower in the CCS region, with only 19% (n=22) reporting extended hours. In addition, 87% (n=627) of all respondents reported that services did not remove other clinical responsibilities to allow the healthcare workers to focus on the extra mpox-related work.

### Prior knowledge, confidence and misdiagnosis

30% (n=218) of respondents reported that they had never heard of mpox before the outbreak; 24% (n=172), 4% (n=32), and 2% (n=14) of those based in Europe, the CCS region and the USA and Canada, respectively. Fewer than 1% (n=6) of respondents indicated that they had seen and treated an mpox case prior to this outbreak.

Amongst respondents in Europe, 60% (n=319) described themselves as not at all or only a little bit confident managing suspected or confirmed clinical cases of mpox at the beginning of the outbreak. In the CCS region, 49% (n=55), and in the USA and Canada 62% (n=50), similarly described themselves as not at all or only a little bit confident. In all the regions, over 25% of the respondents reported that they had misdiagnosed someone with a mpox-related rash with another condition initially. The most common misdiagnoses were chickenpox, syphilis and herpes.

### Training and support

Overall 53% (n=384) agreed or strongly agreed that their institution provided clear, timely and authoritative information about mpox. Agreement with this was lowest in the CCS region at only 31% (n=35), as compared to 56% (n=297) within Europe and 63% (n=52) within the USA and Canada.

59% (n=429) reported that they had received specific education, training or instruction about mpox within their facility. With regards to general outbreak management education, 41% (n=218) of respondents within Europe had completed this training. However, higher proportions were reported in the CCS region and the USA and Canada, 65% (n= 74) and 52% (n=43), respectively.

### Vaccines

Less than 50% of respondents had been vaccinated with the smallpox vaccine before the mpox outbreak. Fewer than one in three of respondents within Europe, the USA and Canada, and fewer than 1 in 10 in the CCS region, had been offered the smallpox vaccine to prevent mpox (ie. as pre-exposure prophylaxis). Once offered, uptake of the vaccine were high across all regions. 71% (n=121), 100% (n=10), and 85% (n=22) accepted and received the vaccine within Europe, the CCS region and the USA and Canada; respectively.

Within Europe, 26% of respondents (n=39) did not feel they received the vaccine in a timely and equitable manner compared to 30% (n=3) and 19% (n=5) in the CCS region, the USA and Canada; respectively. Over 90% of respondents in each region felt vaccination should be offered for mpox to people at high risk of mpox infection prior to exposure, i.e. pre-exposure prophylaxis. However, with regards to vaccination being offered for mpox for all health professionals managing suspected or confirmed clinical cases of mpox, proportions were lower; 61% (n=324) agreed in Europe, 89% (n=101) in the CCS region and 67% (n=55) in the USA and Canada.

Just under half the respondents (n=254) in Europe indicated that vaccination was being offered to all people at high risk in their countries. Comparatively fewer indicated that all people at high risk had been offered vaccination in the USA and Canada (38%; n=31) and only 3% (n=3) in the CCS region. In countries where vaccination was available, 47% (n=340) of HCWs thought access to vaccine for mpox was not at all/slightly adequate in their country; 37% (n=268) in Europe, 3% (n=22) in the CCS region and 7% (n=50) in the USA and Canada.

### Clinical guidelines and national public health agency

The majority of respondents (60%; n=434) reported that their service followed local service guidelines and national guidelines (72%; n=523). Within Europe, only 39% (n=207) reported that their service followed international guidelines as compared to 61% (n=70) in the CCS region and 51.22% (n=42) in the USA and Canada.

When asked about their satisfaction with the support that their service received from their national public health agency, overall 40% (n=284) were not at all or only slightly satisfied with the support. The highest proportion of this dissatisfaction was noted in the CCS region (55%; n=62).

### Moral distress and moral injury

Overall, more than half (55%; n=394) felt that the term ’moral distress’ resonated with their experiences at work managing suspected or confirmed clinical cases of mpox. Over one third (37%; n=265) felt that the term ’moral injury’ resonated with their experiences at work managing suspected or confirmed clinical cases of mpox.

## Discussion

Our multi-national survey study sheds light on the experiences of healthcare workers across the WHO European region and the WHO region of the Americas during the mpox PHEIC of 2022-2023, which unfolded less than 18 months after the second wave of COVID-19 lockdown measures.

Despite mpox being identified in animals 66 years ago and in humans 20 years later in 1970 [9, 10], our study revealed significant gaps in HCW knowledge on this neglected disease. HCWs, predominantly in the fields of HIV, sexual health, and infectious diseases and located outside Africa, showed a concerning lack of clinical familiarity with mpox. According to the survey, 30% of HCWs had never heard of mpox prior to the global outbreaks, and fewer than 1% had encountered and/or treated a case. Of note, in the 2022 epidemic mpox presented in a way that was epidemiologically and clinically distinct from the endemic form of mpox found on the African continent, adding a major challenge to the understanding of this neglected and emerging disease.

This knowledge deficit underscores the urgent need for enhanced global pandemic preparedness initiatives which prioritise HCWs most likely to be at the frontlines of any outbreak response.

A significant proportion (40%) of respondents reported an increased workload, with nearly 90% indicating that their regular duties were not reduced to accommodate the additional burden brought on by the mpox outbreak. This affirms the heightened vulnerability frontline HCWs face, especially when mobilizing a response to a new outbreak [25]. The impact is particularly severe among HCWs with expertise in infectious diseases and related fields, who bear a disproportionate burden in responding to disease outbreaks. Not only did HCWs feel over-burdened in the mpox response, they also felt under-prepared to respond to a new outbreak of a disease with which many of them had little familiarity. This lack of support and training was strongest among respondents in the CCS region. Throughout the COVID-19 pandemic, many frontline workers have experienced increased physical and psychological stressors and burnout, leading some to ultimately leave the workforce [3–5]. Similar patterns during the mpox outbreak emphasise that, despite the well-recognised concern of the risk of burnout and increased workloads during health crises, health systems still fall short in measures to support HCWs. Such disconnect highlights a critical gap in our approach to healthcare preparedness and HCW wellbeing as an important pillar of pandemic readiness. Particularly concerning is the mpox impact on HCWs who deliver HIV, sexual health, and infectious disease services - specialties that in many countries were already grappling with a crisis of attrition in the workforce due to insufficient funding and low compensation in many health economies [26].

In this study, vaccine acceptance and uptake were generally high among HCWs in the sample, yet significant issues of vaccine inequity and access emerged. Participants from Europe and North America indicated moderate access to preventive small pox vaccines for pre-exposure vaccination (35-50%), in stark contrast to those in the CCS region, where access by HCWs fell below 6%. This discrepancy mirrors the ongoing neglect and lack of prioritisation faced by low-resourced countries in Africa, where mpox has been endemic for over five decades [9, 10]. The effective containment of the outbreak in North American and European countries was partly due to accessible vaccines for those most at risk of mpox exposure. Moreover, the acquisition of mpox in healthcare settings has been recurrently documented, particularly in endemic areas. HCWs face heightened infection risks while caring for mpox patients, exacerbated by inadequate resources for proper infection control, such as personal protective equipment and sanitary supplies, alongside overcrowding in healthcare facilities. Thus, vaccinating HCWs, especially during outbreaks in regions with scarce infection prevention resources, becomes crucial and should be prioritised. Our survey was designed to evaluate clinicians respsonses to an unfamiliar pathogen and was therefore disseminated to HCWs who had not dealt with mpox before within their health system. Our survey underlines the importance of conducting similar studies led by African clinicians to assess the access of HCWs to essential tools like vaccines and the impact of such limitations on their perceptions of support, moral distress, and moral injury as they address ongoing and evolving mpox outbreaks in these settings. Building a resilient HCW workforce that is equipped to effectively respond to emerging outbreaks of disease and pandemics also implies prioritising the protection of the workforce with tools and resources which allow them to feel confident and secure in performing their duties.

More than half of our participants reported identifying with the term "moral distress" based on their experiences in managing mpox cases, and a third felt that "moral injury" described their experiences. Although some level of moral distress may be inevitable for healthcare workers during times of increased workload and stress, the high proportion of respondents in our study who reported these feelings is concerning. A manuscript analysing open-text responses to these questions and exploring responses in greater depth is forthcoming [27]. Even when HCWs find ways to be resilient, an excess workload with limited support and resources is unsustainable. Systemic changes are urgently needed to address understaffing, low compensation and lack of proper incentives in routine care delivery, as these are even, more challenging to address in times of crisis. Additionally, efforts must also go towards repairing the damage done to the morally injured HCWs by fostering community among frontline workers who have shared experiences.

A number of limitations of our study should be highlighted to avoid over-interpretation of the results. Firstly, this was an exploratory study that relied on a convenience sampling strategy; thus limiting its generalisability. Secondly, although the demographic breakdown of respondents appears to reflect the regional health care workforce, it may not be truly representative. A significant proportion of respondents were from Europe and this may further limit the global applicability. Thirdly, it focussed on the quantitative data from a cross-sectional survey and an inherent element of this design is recall bias. Respondents were offered to share further perspectives via free text and these responses are reported elsewhere. Fourthly, the sample size meant that the study was underpowered and this restricted the statistical analyses that could be undertaken and the inferences that could be made. This also limited any temporal changes being elicited. Finally, the mpox PHEIC occurred in varying socio-political contexts which likely impacted experiences and perspectives which were not specifically explored with the survey. Further qualitative exploration is needed.

## Conclusion

Our survey study clearly illustrates the harsh realities faced by HCWs facing the global mpox epidemic of 2022-2023 in their health system for the first time during amidst the lingering effects of the COVID- 19 pandemic. It highlights a critical need for addressing the profound gaps in HCW knowledge about re-emerging diseases with pandemic potential like mpox, and underscores the urgency of strengthening the resilience of global health systems and multidisciplinary approaches against future outbreaks. The experiences of HCWs, marred by increased workloads without commensurate support, inadequate access to crucial vaccines, and pervasive feelings of moral distress and injury should serve as a clarion call for immediate action.

As we navigate the aftermath of the global epidemic mpox emergency and anticipate future health crises, it is imperative that we prioritise the well-being and preparedness of those on the front lines. This includes not only equipping HCWs with the necessary knowledge and resources to effectively respond to disease outbreaks, but also ensuring their mental and psychological needs are addressed. These initiatives must include a priority focus on HCWs in Africa’s endemic regions, who have been battling mpox outbreaks for years yet remain overlooked in global response plans. Moving forward, it is crucial that the insights we have gained from the mpox outbreak and the firsthand accounts of healthcare workers pave the way for a renewed approach to pandemic preparedness. This approach must prioritise the health, safety, and respect of those who dedicate their lives to caring for others.

## Data Availability

The protocol, all survey questions examined in this analysis and relevant data are available online at https://osf.io/jf6kw.

## Acknowledgements

Mpox SHARE-NET writing group: Sara Paparini, Chikondi Mwendera, Anthony K J Smith, Omar Sued, Jonathan M Schapiro, Ann-brit Eg Hansen, Carlos Del Rio, Dimie Ogoina, Raquel Martin Iguacel, Elena Sendagorta

